# Factors impacting prescription practice in primary healthcare setting in India: A case study in Rajasthan

**DOI:** 10.1101/2020.06.17.20133439

**Authors:** Arup Kumar Das, Shyama Nagarajan, Ruchi Bhargava, Rajesh Ranjan Singh, Ambey Kumar Srivastava, Amitabh Dutta

**Affiliations:** Lords Education and Health Society (LEHS)| Wadhwani Initiative for Sustainable Healthcare (WISH); SahaManthran Pvt Ltd; Institute of Anaesthesiology, Pain, & Perioperative Medicine; Member Secretary, Ethics Committee, Sir Ganga Ram Hospital, New Delhi

**Keywords:** Prescription practice, adequacy, appropriateness, legibility, compliance, comprehensiveness, primary health care, poly-pharmacy, signs and symptoms

## Abstract

A.

**Background:** We present a case of prescription practices in the Indian state of Rajasthan to demonstrate the effect of provider and system level factors, and their interactions on good prescription practices. We have presented two major dimensions of good prescription practice; first, completeness of prescription, a measure of adequacy; and second, appropriateness of prescription, a measure of quality of care (QoC).

**Method:** We used mixed method to audit 2801 prescriptions, selected from 24 rural and 7 urban government Primary Health Centres (PHCs) of Rajasthan, India. The findings represent analysis of 97% of the ‘selected prescriptions’ that were considered ‘legible’. The ‘selected prescriptions’ ensure represent variance in patient categories, seasonality and number of OPD foot fall across days in a week. Semi-structured in-depth interviews, followed by Focused Group Discussion (FGD) with providers was undertaken to obtain insights about facilitators and barriers to good prescription practices. We compared descriptive statistics across quintiles on adequacy indicators to understand variations around provider and system level factors; multilevel logistic regression model was used to obtain the adjusted effect. To assess appropriateness of quality of care (QoC), we evaluated 783 prescriptions that had adequate information to compare factors impacting QoC across quintiles. Finally, findings from the FGD was used to substantiate findings from the quantitative analysis.

**Result:** We found that prescription practices are outcomes of both provider and system level factors, and their interactions. The documentation of patient complaint, examination findings largely depends on system level factors; 59% and 38%, respectively. The treatment adequacy is largely associated with patient category. Ownership compliance of the doctors, measured in terms of their signature in the prescriptions, emerged as an important factor determining both adequacy and accuracy of prescriptions. We also found that treatment appropriateness, measured in terms of QoC, depends on both provider and system level factors. A conducive environment and trained provider are more likely to provide adequate and appropriate treatment. It is also observed that higher patient load is not a counter-productive to treatment appropriateness. Out of 783 legible prescriptions that were assessed for its appropriateness, only 36% were found inappropriate in terms of their documented justification for the treatment advised.

**Conclusion:** There is a need to focus on provider and system level factors to improve prescription practices in primary health care (PHC). We recommend that institutional strengthening at systemic and provider level using innovative ways; such as task shifting to nurses as ‘physician assistants’, and reducing administrative activities of physicians to enhance focus on clinical work can propel better prescription practice.

## A. Introduction

Rational medicine use requires patients to receive medication appropriate to their clinical needs, in doses that meet their requirements, for an adequate period of time, and at a cost affordable to the community.^1,2,3^ Unfortunately, WHO report, 2002 on rational drug use reports that more than 50% of all medicines prescribed, dispensed, or sold on a global basis are inappropriate; and around 50% of inappropriateness is contributed by patients in terms of their failure to take medicines as prescribed.^4^ Bhatnagar and Mishra have reported that overuse of medicines stimulates inappropriate patient demand and leads to medicine stock-outs and loss of patient confidence in the health system.^5^ Inappropriate (over or underutilization) use of medicines increases antimicrobial resistance, contributes to poor clinical outcomes, and manifests as avoidable adverse drug reactions.^4^ Inappropriate usage is also a wastage of scarce economic resources.^5^ Overall 25%–70% health expenditure in developing countries is contributed by expenditure on medicines, as against 10% in high-income countries.^6^

The situation as regards drug use is no different in India from rest of the world. One third of world’s population living in India lacks access to essential medicine, which makes it up to 65% in India alone.^7, 8^ With out-of-pocket expenditure on health at 82%, 62% is spent on medicines; large percentage of which is rendered waste because of irrational prescription of non-essential medicines, and brands instead of generics.^9^ A study by Kasabi et.al. in Karnataka with 200 prescriptions, in 15 PHCs in a district, reported more than 45% use of antibiotics at primary level.^10^

The prescription practice indicators as defined by WHO is an objective measure to describe patterns of medicine usage, including use of antibiotics, injections, and generic from a defined Essential Drug List (EDL).^11,12^

WHO apart, The Medical Code of Ethics prescribed by the Medical Council of India (MCI) in 2002, amended in 2016 regulates prescription practice in India; to foster rational drug use, and assure completeness of prescribed care plan to ensure patient safety.^13^ The prescription in India is thus a legally admissible evidence to care process, generated by doctors and includes the following details as shown in Table 2:

**Table 1:** WHO prescription practice indicators and recommended reference values

| WHO prescription practice indicators | Reference value |
| --- | --- |
| Average number of medicines per encounter | <2 |
| Percentage of medicines prescribed by generic name | 100 % |
| Percentage of encounters with an antibiotic prescribed | <30 % |
| Percentage of encounters with an injection prescribed | <20 % |
| Percentage of medicines prescribed from an EDL or formulary | 100 % |

**Table 2:** MCI 2002 Guidelines for prescription writing

| s.no | Indicators | Details of indicators as per MCI Guideline—2002 |
| --- | --- | --- |
| 1. | **Patient's demographic profile** | Document the name, age, gender, out-patient registration number to be able to track continuity of care; preferably occupation to draw requisite correlation to disease propensity |
| 2. | **Date of prescription** | To track course of treatment |
| 3. | **Patient findings** | Document chief complaints (presenting signs and symptoms) of the patient, examination findings of the clinician to evaluate patient progress overtime and ensure continuity of care |
| 4. | **Provisional diagnosis made by clinician** | To assist in disease codification, thereby contribute to understanding the disease morbidity patterns and propensity to diseases in the said community |
| 5. | **Investigation advised** | Document investigations to clinch the diagnosis with an evidence (laboratory or imaging or others), and assist in continuum of care and avoid ethical dilemmas during treatment processes |
| 6. | **Medicine prescribed and care plan shared-** | Documentation to guide the care providers (nurses in the hospital and family in a home setting) and patient themselves to support early recuperation and precautions to be taken to avoid future occurrence. This is critical for continuity of care at facility level to avoid errors of commission and/or omission, to assure cooperation of patients and their families in the care process as a team. This section of prescription is considered to be complete, if it includes:<br>• Drugs written in capital/ legible letters with dose,<br>• Mentions total duration over time, route, frequency-number of times per day, and time of administration of drug<br>• Instruction for administering the medication (if applicable—before or after food, among others)<br>• Cautions to watch for potential drug reactions, common side effects among others |
| 7. | **Follow-up and Referral** | The MCI Code of Conduct, 2002 for medical practitioners in its sections 3.5 and 3.6 elaborates on follow-up and referrals. The MCI expects that no decision should restrain the attending physician from making subsequent variations in the treatment during next consultation; provided reasons for the variations are discussed/ explained. Also, when a patient is referred to a specialist by the attending physician, a case summary of the patient should be given to the specialist; and who in-turn should communicate his opinion in writing to the attending physician. |
| 8. | **Validation by clinician that the prescription is written by him/her** | Signing the prescription with name and registration number as a licensed practitioner to justify reasonableness of prescription/ care provided w.r.t the specialization of the prescribing clinician. |

While there are some studies, there are far and few elaborating barriers to good prescription practice, and its correlation to confounding factors. Therefore, this study was intended to pave way for understanding challenges in prescription practices in primary care level and draw possible solutions to sustain a good prescription practice in a PHC setting.

Under this backdrop, the Wadhwani Initiative for Sustainable Healthcare (WISH), a not-for-profit initiative of the Lords Education &Health Society (LEHS), conducted the present study in Rajasthan. Therefore, we defined and designed the study to identify factors associated with adequacy and appropriateness of prescribing practices at primary level.

- ‘Adequacy’ means, compliance to completeness and comprehensiveness of prescription practices as per WHO norms and MCI Guidelines;
- ‘Appropriateness’ means, the extent to which the practices can be justified in terms of ‘reasonableness’ of quality of care (QoC).^14^ Reasonableness in QoC stems from variation in qualification (knowledge gained as a medical graduate or postgraduate in specific speciality) and experience (years of relevant patient care service provided).

## B. Methods

A sequential mixed-method approach was adopted to evaluate the prescribing practices in the 31 PHCs in 11 districts of Rajasthan as shown in the flow diagram in Figure-1. Prescriptions for a period of two years from September 2016-August 2018 were reviewed. In-depth interviews (IDI) with PHCs’ service providers were conducted to capture their profile. Subsequently a focused group discussion (FGD) was held to share findings of the quantitative study, and deliberate on barriers and facilitators to good prescription practice, and develop solutions to improve practice.

**Figure 1:**
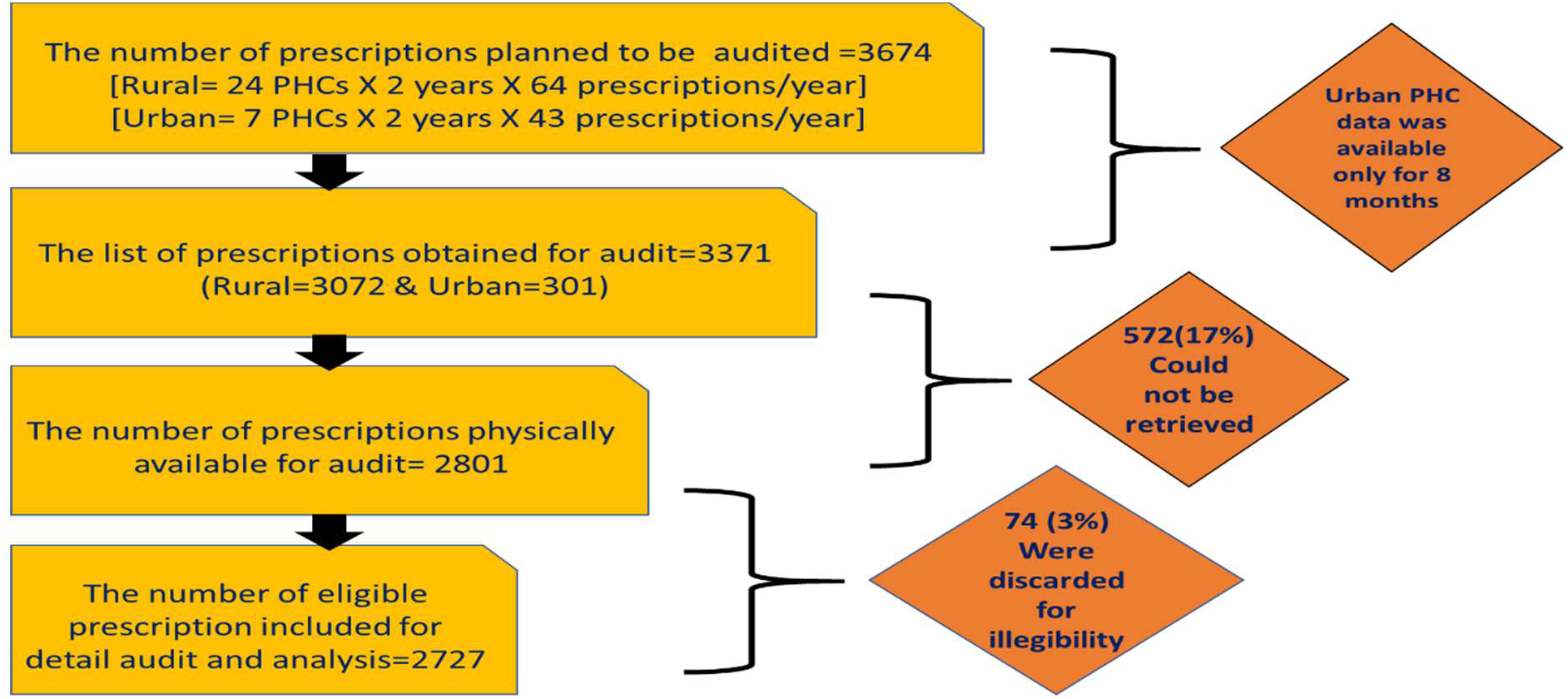
Targeted vs actual number of prescriptions included in the study

A total of 3698 prescriptions **@64 prescriptions per year from 24 rural PHCs each and 43 per year from 7 urban PHCs each were selected**. As urban PHCs got operationalised from January 2018, prescriptions during the 8-month period of their operationalization could be collected. Out of 3698, 572 prescriptions (17%) could not be retrieved because of loss of selected prescriptions in constrained storage conditions of PHCs that were operating out of make shift buildings. Of the total sample of 2801 prescriptions, 2727 (97%) prescriptions were defined as legible for further analysis and remaining 3(%) were discarded as illegible.7

While selecting the prescriptions, considerations were made to ensure comprehensive inclusion of heterogeneity with respect to patient category, seasonality, and variations in out-patient (OP) footfall across the days in a week at the PHCs.

**To ensure adequate representation of profile of diseases being treated across patient categories**, we picked samples from all five different patient categories (Table 2) (i) maternal (M), women who availed maternity services, (ii) paediatrics (P), sub-divided into under-five(P1) and children between 5-15 years (P2); (iii) Adults (A), all cases other than maternity; (iv) old age over 60yrs (O); and (v) emergency (E) cases, to capture all emergencies and trauma.^8^

**To capture seasonal variation of disease profile**, calendar year was split into four quarters (i) Winter- November to January, (ii) Spring-February to April, (iii) Summer—May to July, (iv) and Rainy—August to October. Further, it was ensured that all six working days of a week has equal probability of being selected twice every season, as shown in Table 4.^9^

**Table 3:**
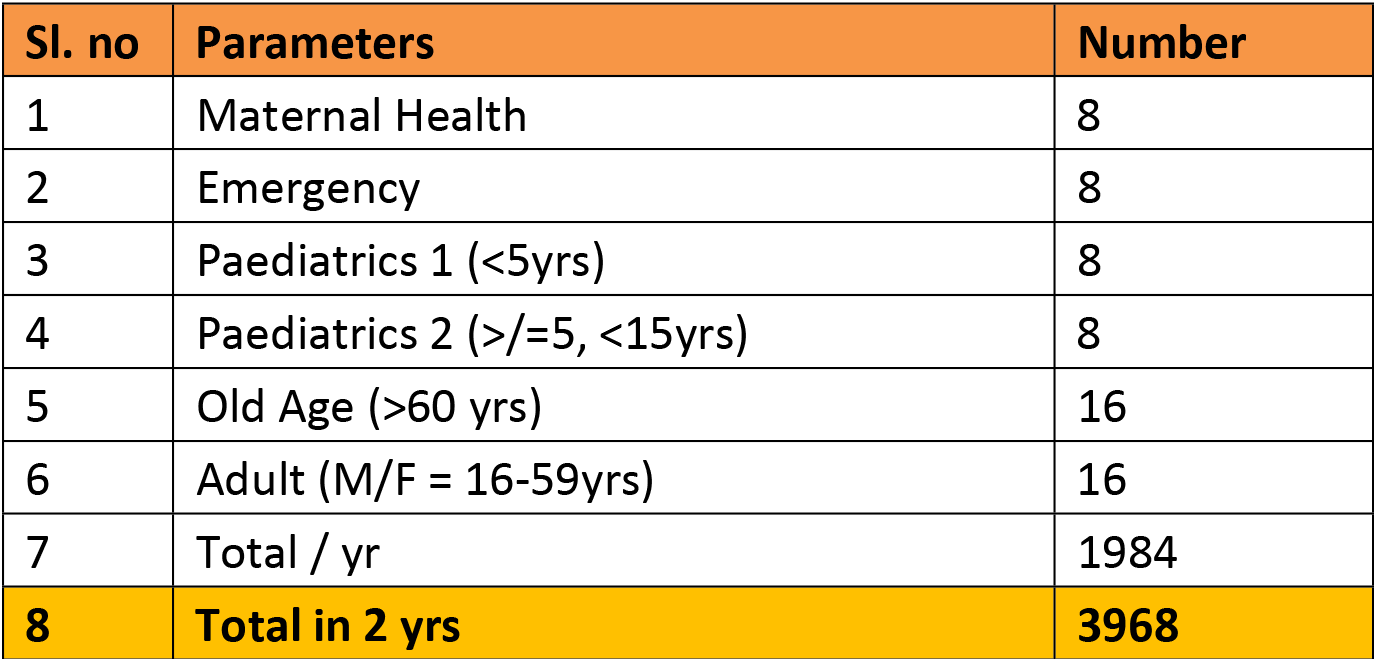
Estimated sample according to patient profile across 31 PHCs

| Sl. no | Parameters | Number |
| --- | --- | --- |
| 1 | Maternal Health | 8 |
| 2 | Emergency | 8 |
| 3 | Paediatrics 1 (<5yrs) | 8 |
| 4 | Paediatrics 2 (>=5, <15yrs) | 8 |
| 5 | Old Age (>60 yrs) | 16 |
| 6 | Adult (M/F = 16-59yrs) | 16 |
| 7 | Total / yr | 1984 |
| 8 | **Total in 2 yrs** | **3968** |

**Table 4:** Allocation of sample prescriptions according to patient category, week days and four seasons

| Months | Nov and May |  |  |  | Dec and Jun |  |  |  | Jan and Jul |  |  |  |
| --- | --- | --- | --- | --- | --- | --- | --- | --- | --- | --- | --- | --- |
| Days | M | W | F | SA | M | TU | TH | SA | TU | W | TH | F |
| Specialties | M/E | P1/P2 | O | A | P1/P2 | O | A | M/E | O | A | M/E | P1/P2 |
| Samples to be collected/ month | 2 | 2 | 4 | 4 | 2 | 4 | 4 | 2 | 4 | 4 | 2 | 2 |
| Months | Feb and Aug |  |  |  | Mar and Sep |  |  |  | Apr and Oct |  |  |  |
| Days | M | W | F | SA | M | TU | TH | SA | TU | W | TH | F |
| Specialties | A | M/E | P1/P2 | O | M/E | P1/P2 | O | A | P1/P2 | O | A | M/E |
| Sample to be collected/ month | 4 | 2 | 2 | 4 | 2 | 2 | 4 | 4 | 2 | 4 | 4 | 2 |

To avoid investigator bias, serial number wise first occurrence/s for every patient category in the OP register was identified to collect desired number (2-4) of prescriptions.

Semi-structured interviews were conducted to obtain profile the providers (one doctor, one nurse and one pharmacist per facility). In all we could interview 23 doctors, 31 nurses and 28 pharmacists based on their availability during the study period. At the final stage 15 medical officers, 6 GNMs/pharmacists participated in the FGD.

The quantitative analysis was aimed at estimating the percentage compliance to WHO indicators and MCI requirements as follows:

a. ***Completeness of prescription—***This was assessed on the seven elements of completion, as per the MCI guidelines.
  - **Demographic compliance**--% of Prescription with complete demographic information; such as Name, Age, Sex, Date of visit, Contact Details.
  - **Registration process compliance**--% of Prescription with details on unique id to support identification of patient throughout the care process.
  - **Patient compliant compliance**--% of Prescription with complete in terms of recording of signs, symptoms presented by the patients.
  - **Doctor examination compliance**--% of Prescription with complete recording of weight, BP, Provisional Diagnosis, and investigation ordered.
  - **Treatment compliance**--% of Prescription with complete recording of final diagnosis, prescription of medication (completeness refers to information on Name of the drug, dose, strength and Number of days to be consumed) and care plan. This also includes % of prescription with Drugs written in capital letters as per MCI requirement
  - **Continuum of care compliance**--% of Prescription with complete recording of requisite conditions for a follow-up, and referral
  - **Ownership compliance**--% of Prescription with complete signature of the treating clinician.
b. ***Appropriateness of prescription***—refers to analysis of prescription on quality of care in terms of diagnosis, investigation and treatment prescribed. Appropriateness was analysed at two levels:
  - **Facility level** to understand the systemic issues impacting appropriateness;
  - **Clinical level** to understand the clinical issues as regards patients’ complaints, doctor’s findings, investigation advised, care plan designed and treatment offered.

We have used bi-variate and multivariate analysis to correlate factors associated with good prescription practices. Multilevel logistic analysis, random slope model was used to understand the role of facility or provider level factors affecting prescription practices. The model comparison was carried out using Akaike Information Criteria (AIC) and Bayesian Information Criteria (BIC), a lower value of these statistic indicates best fit model.

## C. Results

The findings of the study as revealed were:

### 1. Completeness of prescription

Completeness evaluated on seven broad sections as under.

a. ***Demographic Details*:** The *first compliance* was documenting of basic demographics in the prescription; meaning noting of name, age, gender, address and contact details of the patients. While name and age of patient was recorded in all eligible prescriptions; sex was not recorded in 65 (2.3%) prescriptions. Highest compliance level was seen in prescriptions for elderly (29%); and lowest being that for maternity and emergency cases, at 5%. While MCI mandates capturing of data on occupation and place of living, the same was not captured in any of the audited prescriptions. These socio-economic indicators have significant bearing on the disease manifestation, its recurrence/ chronicity. For instance, recurrence of dermatitis and staining of tooth in some PHCs was attributed to high floride content in ground water by the staff. Repeated treatment of such conditions without documentation of living condition, reduces the probability of escalating issues with relevant state authorities to systematically address the public health hazard.
b. ***Registration Process Details:*** *The second compliance*, includes assigning registration number to each patient and recording visit dates, to track patient progress during follow ups, and referrals to other/specialised services. The study reveals that while assigning registration number was almost universal; there was no inter or intra facility standard followed to assign registration numbers. Each year, the registration number began with serial number ‘1’ and continued; thereby creating new registration numbers for the same patient on each visit. In the absence of standard numbering process, a mechanism to identify new and repeat/ follow-up cases doesn’t exist; except for the informal facial recognition by the hospital staff. Audited prescriptions revealed 1- 8% duplication of registration numbers in 8 of 31 facilities; except for one PHC that had 56% duplication. During FGD lack of documentation of dates was cited as cause for duplication of registration numbers. Overall, 16% of the eligible prescriptions did not have the date recorded, ranging from zero to 100% across PHCs.
c. ***Patient complaint details*:** *The third compliance is towards recording of patient complaints*. Herein patient’s narrative of chief complaints (symptoms) and provider’s interpretation of these complaints (signs) gets recorded. These signs and symptoms with clinical examination findings as noted by the doctor, enables the clinician to provisionally diagnose the disease; and decide on the investigations required for the final diagnosis, design the treatment &care plan and decide referral, if required. Our study revealed that signs &symptoms were recorded in only 28% of eligible prescriptions. A substantial difference was noted in recording symptoms across 31 PHCs, ranging from zero to 100%, with SD 28%, skewed (0.696) &leptokurtic (2.28) distribution. It was noted that signs and symptoms were used synonymously, and handwriting was hard to decipher in all prescriptions, as shown in Table-5. The terminology used for writing signs and symptoms were either incomplete or not adhered to as per the prescribed textbooks. For instance, itching or skin allergy, was mentioned in 2.2 % cases as ‘skin’. This was attributed to the documentation of prescriptions by the nurses or paramedics; who while were untrained and ineligible, substituted in absentia of the doctor, to respond to patient needs. Among the eligible prescriptions, majority had only one symptom recorded (19.7%) and 8.6% had more than one symptom recorded. Most frequently mentioned signs and symptoms were fever (14.7%), pain (4.7%), diarrhoea (4.1%), injury (3%), pregnancy and delivery (2.2 %). Interestingly, a clustering of symptoms around patient categories was recorded. For example, fever, diarrhoea and skin allergy was clustered around paediatric(P2), adults and old age population. Based on proportion of prescriptions that had recorded signs &symptoms, we classified the PHCs into five quintiles (Table 6) to compare their profiles with provider and facility level factors.^10^ No substantial difference in patient profile in these PHCs, except that the facilities under high quintile had slightly higher load of maternity cases. However, substantial difference was noted across five quintiles in terms of provider and facility level factors. The cluster of 6 PHCs, that had high recording of symptoms (76 %) had providers with longer work experience with our PHCs, had relatively infrequent drug stock-out, more accountable providers (measured on proportion of prescription signed), with higher provider focus on service delivery as against administrative work. **Contrary to the common belief, high OPD load was not a hindrance to recording signs and symptoms**. This was corroborated during the FGD, and doctors recommended reducing their administrative work that consumed 50% of their time, and training AYUSH^11^ practitioners and GNMs for documenting findings in the prescription, to save doctor’s time for patient examination and writing the care plan. A multilevel logistic model was found best fit in comparison to simple logistic regression, which explains 59.5% of the variations attributable to system level factors (Table-7). Further, the emergency cases, paediatric and old age were 2.6 (1.6 – 4.0), 1.5 (0.9 – 2.15) and 1.5 (1.1 – 2.3) times more likely to have their symptoms recorded, respectively than the maternity cases. Qualitative finding revealed that protocoled treatment for maternity cases rendered documentation of signs and symptoms redundant. However, in others, such as in old age and paediatric patients, non-documentation of signs and symptoms had the potential for medication errors; and therefore, compliance was necessitated and hence was high.
d. ***Examination Finding Details:*** The *fourth* compliance on recording of *doctor’s examination findings*, includes noting of patient vitals (Blood Pressure-BP, pulse, weight), investigations required, and provisional diagnosis. The compliance was low across facilities and was limited only to maternity cases. BP was recorded in 4 % of eligible prescriptions and 18 % of maternity cases. Weight and investigations were recorded in 2% and 6% of the total, 11% and 23% of maternity cases, respectively. The major investigations documented for maternity cases were Haemoglobin (15 %), HIV/ELISA (6%), Random Blood Sugar (5%), Urine albumin and STI (4% in each), which were a part of protocoled treatment trained to follow. Provisional diagnosis was recorded in 24 % of total and 86% of maternity cases. Three logistic regression model has been compared to identify the effect of recording symptom &investigations, their interaction on reported diagnosis and a multilevel model to identify the role of system level variations. It was evident from the best fit multilevel model that almost 39% variations in reporting diagnosis can be attributed to system level factors. The fixed part of the model revealed that diagnosis was more likely to be written in maternal health prescriptions. The predicted probabilities of interaction suggest that a prescription that had symptom &signs recorded and investigations/ tests prescribed, were 8 and 5 times, had higher likelihood of documented diagnosis respectively (Table-8). During the FGD, providers iterated on the non-essentiality of recording vitals for all clinically stable patients; except for old-age patients who could potentially deteriorate quickly and maternity cases that had protocol to monitor progress.
e. ***Treatment:*** The *fifth compliance* towards *treatment* dealt with polypharmacy, prescription of antibiotic and injections. The mean polypharmacy rate was 2.8 per prescription, median being 3; slightly higher than the WHO recommended reference value. Overall, 63% of eligible prescription had more than 2 drugs prescribed; highest recorded among the old age at 75% with a mean of 3.15 per prescription; and lowest in maternity cases (51%) with mean 2.6 per prescription. Polypharmacy in maternity was attributed to prescription of nutrients, such as iron and folic acid. Across PHCs, polypharmacy varied from 24% to 80%. As for antibiotics in prescription, 63% of the patient encounters had an antibiotic prescribed, which was double the WHO reference value (<30 %); and 9% had more than one antibiotic. The prescription of antibiotics was highest among paediatric (P1-77 %) and lowest in maternity (31%). Since maternity is not a disease, such a high volume of antibiotic usage needs further investigation. Urban PHCs had more antibiotics (68%) per prescription, than in the Rural (62%). Injection prescription also varied across PHCs, ranging from 0 to 46%, with an average of 15%, within WHO reference indicator. However, prescriptions for maternity and emergency cases had higher proportion of injection/s prescribed, with 23% and 25% respectively. This was attributed to the protocoled routine immunization of pregnant women with TT, and injections for pain relief during emergency. Unlike antibiotics, injection prescribed was substantially low for Urban PHCs (5%) vis-à-vis rural (16%). Multilevel logistic model (table-9) revealed that polypharmacy and prescription of antibiotic depends largely on patient’s profile. An old patient was 3.3 times more likely to experience polypharmacy than others; owing to increased multivitamin prescription among elderly. However, 8-9 times increase in polypharmacy in paediatric patients was linked to use of antibiotics. Interestingly provider or system level factors did not contribute to polypharmacy (8%) and increased antibiotic prescription (7%). However, in case of injections, the system level factors accounted for 14% variation. As for provider level factors, patients that had symptom/s recorded had 1.6 times more likely to receive an injection. Second generation aminoglycosides or broad-spectrum antibiotics were the most commonly prescribed antibiotics in the encounters wherein antibiotics were prescribed. Amoxicillin was prescribed in 28% of encounters, Ciprofloxacin on 17%, and Cephalexin was the least prescribed antibiotic. The other most frequently prescribed drugs were Analgesics, Anti-allergens, Multivitamins, Gastroenterology and respiratory drugs. As regards documentation of drug related information, 99.9% drugs were written by their generic names from the Essential Drug List (EDL) as approved by the state; 21 % drugs were indicated with their strength; 79% with dose; and 92% was documented with duration of treatment. During the qualitative study, doctors explained that documentation of drug strength was irrelevant, because of availability of limited strength/s of drugs at the PHCs.
f. ***Continuum of care (follow up and referral):*** Referral and follow up, the two major components of primary care, had poor compliance across all facilities at 1%. While all the respondents during the qualitative study agreed on the importance of documenting follow up and referral to ensure continuum of care; the reasons for abysmal documentation was attributed to:
  a. The practice of dispensing medicine for 3 days forcing patient’s revisit within 72 hours for a follow-up. Providers explained that in a low literacy setting, it’s important to impress upon the need &incentivise follow up, rather than documenting it; because patients hardly refer to prescription for compliance.
  b. Inadequate space in the existing prescription format
  c. Self-motivation of the chronic patients (TB, typhoid, diabetes, dermatitis etc.)
  d. Disincentive for providers to prescribe referral, owing to absence of a system at higher centres to entertain referrals from PHCs.
g. **Ownership compliance:** Ownership compliance was measured by the proportion of prescription wherein doctor’s signature was documented. Overall 58% prescriptions had documented doctor’s signature; higher in urban (78%) against rural (56%) PHCs (Figure-2). Contrary to the common belief that high patient footfall leads to poor completeness of prescription; a high level of correlation was witnessed between proportion of prescriptions signed by the doctors and its completeness (Figure-3). The several logistic regressions drawn to understand factors associated to ownership compliance revealed that the system level factors accounted for 30% of the variations in the best fit model. Prescriptions at urban PHCs had 5 times more potential of being signed by a doctor than a rural one. Similarly, maternity cases had 1.5–2 times higher likelihood of receiving signed prescription. The logistic regression model suggests that provider’s age, experience, availability of medicine and ability of the clinicians to accord time for clinical practice enhances ownership compliance (table-10). The FGD corroborated our quantitative findings that lesser the doctor’s engagement in administrative activities, greater was focus on clinical care and completeness of prescription. Doctor’s opined that nurses be allowed to sign protocoled routine care, if administrative role for doctors was a system level necessity.

**Table 5:** Classification of signs and symptoms as documented

| Symptom Group (Computed) | Symptoms as recorded in the prescription |
| --- | --- |
| Fever-cold | fever, chill, pyrexia- URI, cough and blood, cold, cough, ART, URTI |
| Typhoid | typhoid |
| Injury | injury, accident injury, head injury, burn, cut, injury/ cut, wound, abscess, skin inflammation, swelling, dressing, swelling on head, pus |
| Diarrhoea & Dehydration | diarrhoea, loose motion, loose motions, dehydration, de-hydration, abdomen pain, vomiting, amoebic dysentery, nausea |
| Headache | headache, vertigo |
| Weakness | weakness, anaraxia |
| Skin Infection | skin infection, fungal infection, allergy, boils, dermic anacus, dermatitis, dermal |
| Pain | pain, body pain, bodyache, back pain, joint pain, pain in limbs, pain in legs, muscle ache, shoulder pain, knee pain, arthritis, foot pain |
| Pregnancy & Delivery | delivery, post delivery, labor pain, miscarriage, aborted foetus, MCH delivery, before and after delivery, ANC, pregnancy, membrane rupture, bleeding PV, hemorrhage |
| Animal Bite | dog bite, monkey bite, scabies |
| Hypertension | hypertension, dizziness, HTN |
| Hb | HB, malnutrition, RBS, BL, weight |
| ENT | eye problem, ear problem, mumps, throat problem, right eye catract |
| Gastro | acidity, constipation, ulcer, gastric, liver pain |
| Diabetes Mellitus | Sugar, DM |
| Urine Infection | urine infection, burning, burning feeling, yellow urine, burning micturation, urine4-5 timesat night |
| Breathlessness | asthama, bronchitis, breathlessness |
| Irregular Menstratution | irregular menstruation, bleeding, PVOC, excessive bleeding, pain lower abdomen, amenorrhea, vaginal discharge |
| Old Age | amnesia |

**Note:** *The signs and symtoms have been noted as it appears in the prescription. However, spellings have been corrected to enable easy reading*

**Table 6:** Reasons for Non-Compliance to patient complaint details as per five quintiles

| Factors associated | v.low<br>(N=7) | low<br>(N=6) | medium<br>(N=6) | High<br>(N=6) | v.high<br>(N=6) | Total<br>(N=31) |
| --- | --- | --- | --- | --- | --- | --- |
| % of prescription, Symptom recorded | 1.3 | 7.9 | 22.5 | 46.1 | 76.3 | 28.3 |
| **Patient level** |  |  |  |  |  |  |
| Maternal | 5.9 | 8.6 | 14.2 | 14.7 | 13.6 | 11.3 |
| Emergency | 9.4 | 7.4 | 8.7 | 9.7 | 6.7 | 8.7 |
| Pediatric <5 yrs | 16.4 | 14.2 | 19.1 | 18.4 | 18.4 | 17.5 |
| Pediatric 5-15 yrs | 18.3 | 18.4 | 16.9 | 19.4 | 20.8 | 18.6 |
| Adult (16-59 yrs) | 28.1 | 32.1 | 20.4 | 20.1 | 22.7 | 24.2 |
| Old age (>60 yrs) | 21.8 | 19.3 | 20.7 | 17.6 | 17.7 | 19.7 |
| **Provider level** |  |  |  |  |  |  |
| Mean age of doctors | 42.4 | 52.4 | 48.9 | 50.7 | 50.4 | 47.8 |
| Age of GNM is more than median age (27 years+) | 36.8 | 61.3 | 35.1 | 21.1 | 55.9 | 39.5 |
| Mean working experience (in Months) | 31.7 | 63.1 | 37.5 | 51.1 | 62.0 | 46.5 |
| Work experience of GNM (35+ months) | 52.2 | 77.7 | 88.7 | 76 | 82.7 | 73 |
| **System level** |  |  |  |  |  |  |
| % Rural | 94.7 | 89.3 | 82.6 | 96.6 | 88.5 | 91.1 |
| Mean OPD per day | 46.1 | 48.6 | 47.2 | 40.3 | 48.6 | 45.7 |
| Providers spending 50+% times in admin work | 62.8 | 50.6 | 0 | 0 | 20.7 | 28.3 |
| Medicine was available | 15.9 | 10.7 | 55.2 | 76 | 85.1 | 46.7 |
| Stock out in past 6 months | 20.5 | 72.4 | 23.8 | 23.2 | 11.5 | 28.2 |
| % of prescription doctor signed | 59.1 | 49.7 | 51.6 | 62.8 | 63.9 | 57.9 |
| # of prescriptions (n) | 761 | 413 | 522 | 621 | 410 | 2,727 |

**Table 7:** Role of system level factors on recording signs and symptoms

|  | Odds Ratio | P>z | [95% Conf. Interval] |  | Odds Ratio | P>z | [95% Conf. Interval] |  |
| --- | --- | --- | --- | --- | --- | --- | --- | --- |
| Model 1: Logistic |  |  |  |  | Model 2: Multilevel logistic |  |  |  |
| Fixed part |  |  |  |  |  |  |  |  |
| Patient category |  |  |  |  |  |  |  |  |
| Maternal (ref) | 1.00 |  |  |  | 1.00 |  |  |  |
| Emergency | 1.17 | 0.40 | 0.81 | 1.68 | 2.56 | 0.00 | 1.61 | 4.06 |
| Pediatric <5 yrs | 0.92 | 0.60 | 0.67 | 1.26 | 1.29 | 0.20 | 0.87 | 1.90 |
| Pediatric 5-15 yrs | 1.01 | 0.96 | 0.74 | 1.38 | 1.46 | 0.05 | 1.00 | 2.15 |
| Adult (16-59 yrs) | 0.80 | 0.14 | 0.59 | 1.08 | 1.34 | 0.13 | 0.92 | 1.96 |
| Old age(>60 yrs) | 0.87 | 0.37 | 0.64 | 1.18 | 1.55 | 0.03 | 1.05 | 2.28 |
| Location |  |  |  |  |  |  |  |  |
| Rural (ref) | 1.00 |  |  |  | 1.00 |  |  |  |
| Urban | 1.06 | 0.68 | 0.79 | 1.42 | 1.12 | 0.91 | 0.17 | 7.14 |
| _cons | 0.42 | 0.00 | 0.33 | 0.54 | 0.16 | 0.00 | 0.06 | 0.39 |
| Random part |  |  |  |  |  |  |  |  |
| var(_cons) |  |  |  |  | 4.82 |  | 2.71 | 8.60 |
| Model Summary |  |  |  |  |  |  |  |  |
| ICC |  |  |  |  | 0.59 |  | 0.45 | 0.72 |
| AIC | 3254.55 |  |  |  | 2276.46 |  |  |  |
| BIC | 3295.92 |  |  |  | 2323.75 |  |  |  |

**Table 8:** Facility level factors impacting compliance to documentation of examination findings

| Diagnosis | M1: Logistic |  |  | M2: Logistic with interaction |  |  | M3: Multilevel logistic with interactions |  |  |
| --- | --- | --- | --- | --- | --- | --- | --- | --- | --- |
|  | Odds Ratio | P>z | [95% Conf. Interval] | Odds Ratio | P>z | [95% Conf. Interval] | Odds | P>z | [95% Conf. Interval] |
| **Fixed part** |  |  |  |  |  |  |  |  |  |
| **Patient category** |  |  |  |  |  |  |  |  |  |
| Maternal | 1.000 |  |  | 1.000 |  |  | 1.000 |  |  |
| Emergency | 0.054 | 0.000 | 0.034 0.085 | 0.060 | 0.000 | 0.037 0.095 | 0.042 | 0.000 | 0.024 0.072 |
| Pediatric <5 yrs | 0.019 | 0.000 | 0.012 0.030 | 0.021 | 0.000 | 0.013 0.033 | 0.007 | 0.000 | 0.004 0.013 |
| Pediatric 5-15 yrs | 0.014 | 0.000 | 0.009 0.021 | 0.014 | 0.000 | 0.009 0.023 | 0.005 | 0.000 | 0.003 0.008 |
| Adult (16-59 yrs) | 0.017 | 0.000 | 0.011 0.026 | 0.018 | 0.000 | 0.012 0.028 | 0.008 | 0.000 | 0.005 0.014 |
| Old age(>60 yrs) | 0.017 | 0.000 | 0.011 0.027 | 0.019 | 0.000 | 0.012 0.029 | 0.008 | 0.000 | 0.004 0.014 |
| **Symptom recorded** |  |  |  |  |  |  |  |  |  |
| no | 1.000 |  |  | 1.000 |  |  | 1.000 |  |  |
| yes | 8.418 | 0.000 | 6.634 10.683 | 9.894 | 0.000 | 7.729 12.666 | 7.709 | 0.000 | 5.516 10.772 |
| **Test recorded** |  |  |  |  |  |  |  |  |  |
| no | 1.000 |  |  | 1.000 |  |  | 1.000 |  |  |
| yes | 1.537 | 0.041 | 1.017 2.323 | 6.624 | 0.000 | 3.454 12.706 | 5.285 | 0.000 | 2.482 11.256 |
| **Symptom X Test** |  |  |  |  |  |  |  |  |  |
| yes#yes |  |  |  | 0.097 | 0.000 | 0.042 0.227 | 0.169 | 0.000 | 0.062 0.458 |
| _cons | 4.075 | 0.000 | 2.895 5.735 | 3.484 | 0.000 | 2.476 4.903 | 4.996 | 0.000 | 2.581 9.671 |
| **Random Part** |  |  |  |  |  |  |  |  |  |
| var(_cons) |  |  |  |  |  |  | 2.067 |  | 1.163 3.675 |
| ICC |  |  |  |  |  |  | 0.386 |  | 0.261 0.528 |
| **Model Comparison** |  |  |  |  |  |  |  |  |  |
| AIC | 2037.676 |  |  | 2010.757 |  |  | 1580.959 |  |  |
| BIC | 2084.964 |  |  | 2063.955 |  |  | 1640.069 |  |  |
| BIC | 2084.964 |  |  | 2063.955 |  |  | 1640.069 |  |  |

**Table 9:**
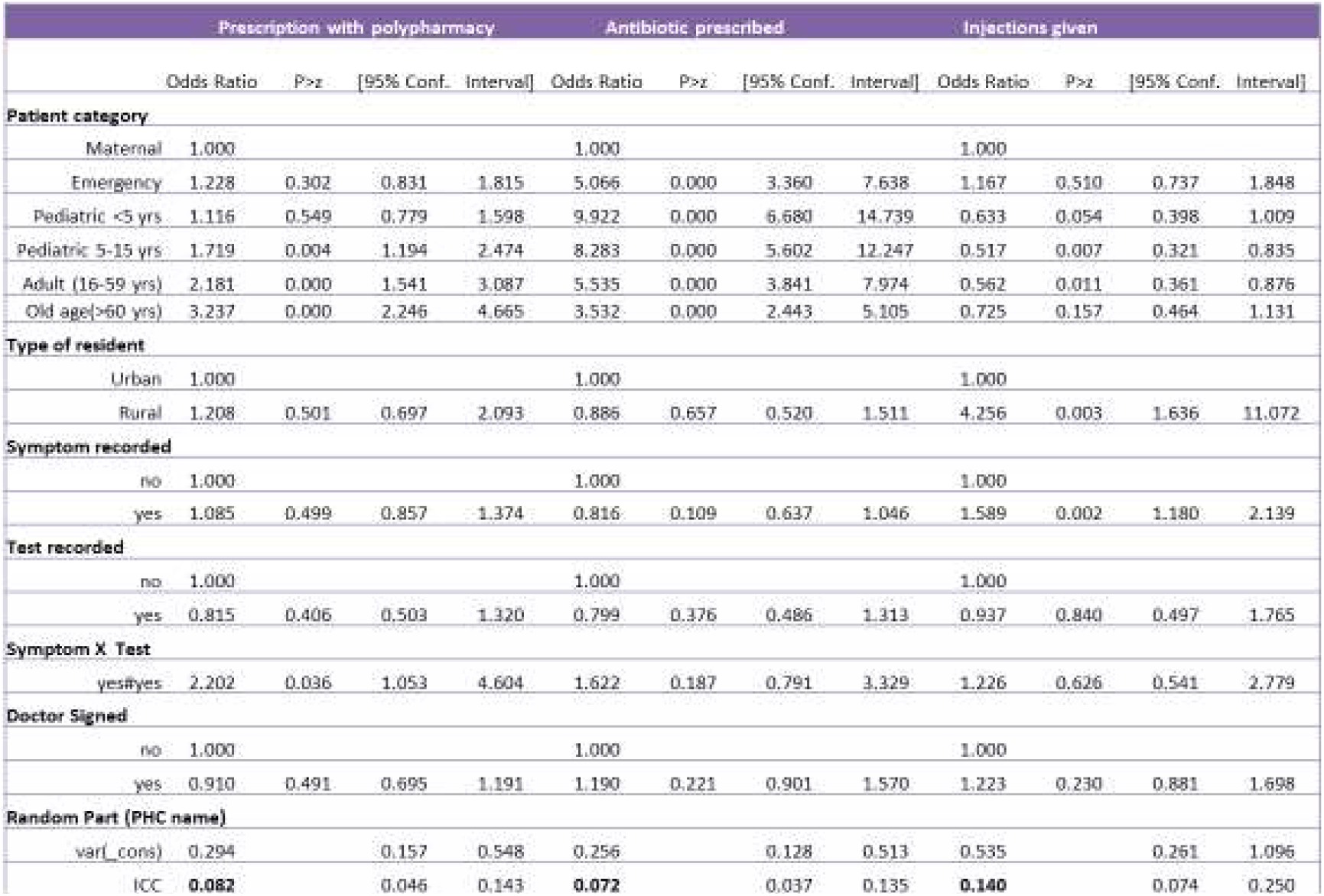
Factors associated with treatment compliance

**Figure 2:**
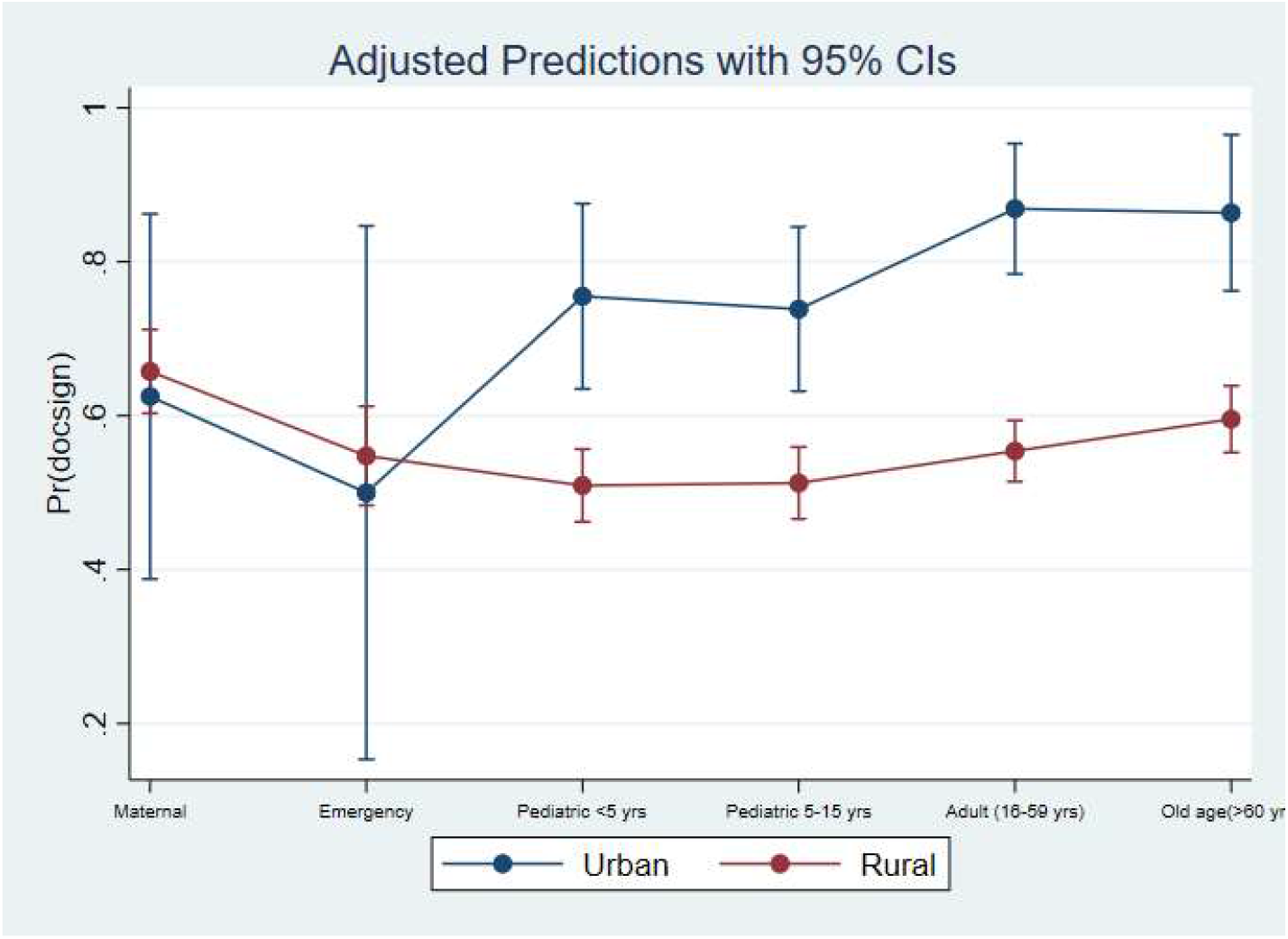
Predicted probability of prescriptions signed by the doctors by patient category and facility location

**Figure 3:**
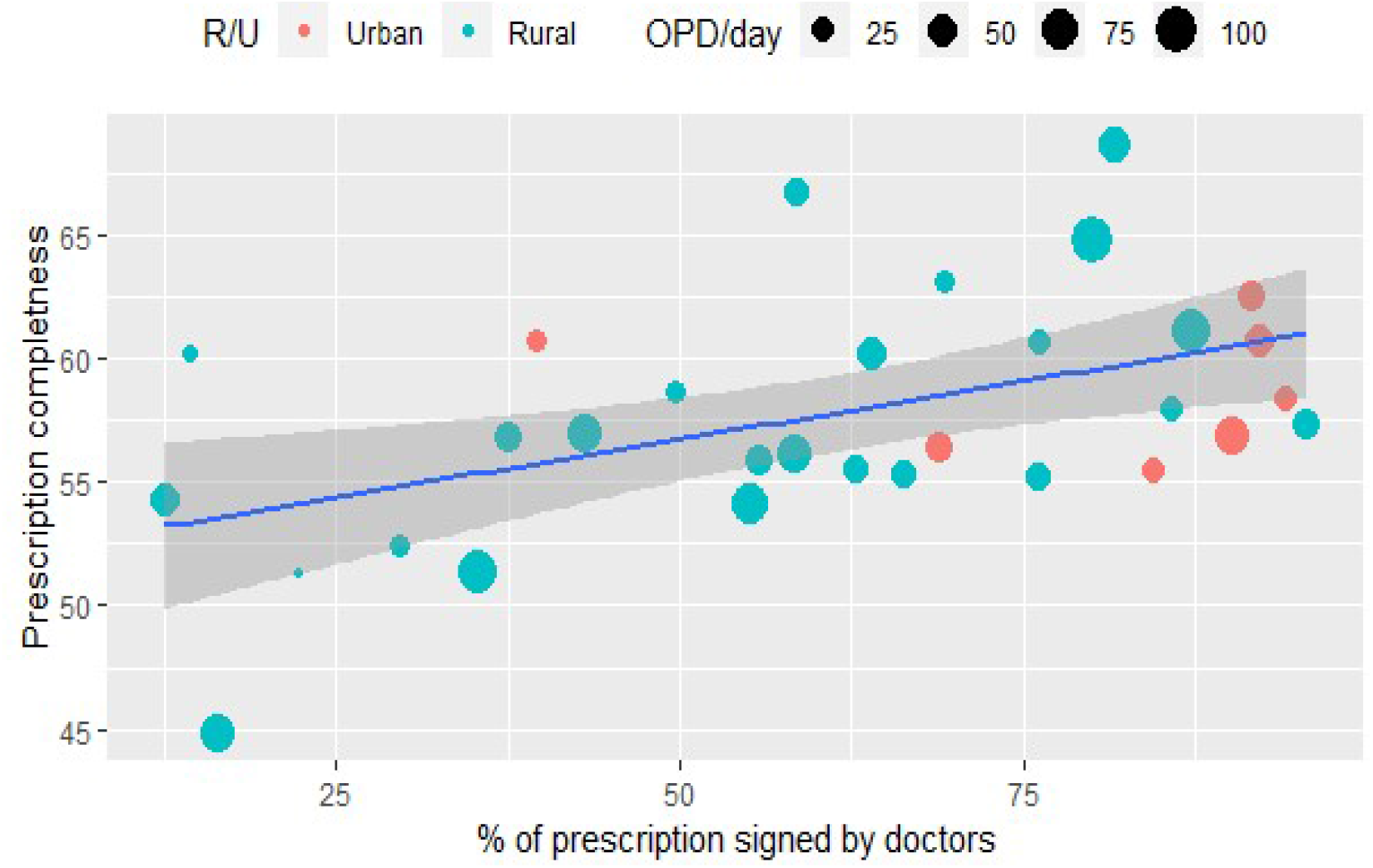
Scatter plot of completeness of prescription and ownership by OPD load and facility location

**Table 10:** Factor influencing ownership compliance

| Doctor Signed | Model-I |  |  |  | Model-II |  |  |  | Model-III |  |  |  |
| --- | --- | --- | --- | --- | --- | --- | --- | --- | --- | --- | --- | --- |
|  | Odds Ratio | P>z | [95% Conf. Interval] |  | Odds Ratio | P>z | [95% Conf. Interval] |  | Odds Ratio | P>z | [95% Conf. Interval] |  |
| Fixed Part |  |  |  |  |  |  |  |  |  |  |  |  |
| Facility location |  |  |  |  |  |  |  |  |  |  |  |  |
| Urban | 1.000 |  |  |  | 1.000 |  |  |  | 1.000 |  |  |  |
| Rural | 0.539 | 0.003 | 0.360 | 0.808 | 0.294 | 0.105 | 0.067 | 1.292 | 0.197 | 0.004 | 0.066 | 0.589 |
| Patient category |  |  |  |  |  |  |  |  |  |  |  |  |
| Maternal | 1.000 |  |  |  | 1.000 |  |  |  | 1.000 |  |  |  |
| Emergency | 0.790 | 0.204 | 0.549 | 1.137 | 0.698 | 0.078 | 0.469 | 1.041 | 0.696 | 0.075 | 0.467 | 1.037 |
| Pediatric <5 yrs | 0.674 | 0.013 | 0.493 | 0.920 | 0.578 | 0.002 | 0.410 | 0.814 | 0.574 | 0.002 | 0.408 | 0.809 |
| Pediatric 5-15 yrs | 0.661 | 0.009 | 0.485 | 0.902 | 0.548 | 0.001 | 0.389 | 0.772 | 0.545 | 0.000 | 0.387 | 0.767 |
| Adult (16-59 yrs) | 0.765 | 0.077 | 0.568 | 1.030 | 0.641 | 0.008 | 0.461 | 0.891 | 0.638 | 0.008 | 0.459 | 0.887 |
| Old age(>60 yrs) | 0.932 | 0.654 | 0.685 | 1.268 | 0.748 | 0.096 | 0.532 | 1.052 | 0.744 | 0.090 | 0.529 | 1.047 |
| Doctors Age |  |  |  |  |  |  |  |  |  |  |  |  |
| <35 years | 1.000 |  |  |  | 1.000 |  |  |  |  |  |  |  |
| 35-65 years | 1.378 | 0.139 | 0.901 | 2.106 | 0.948 | 0.952 | 0.167 | 5.375 |  |  |  |  |
| 65+ years | 1.393 | 0.029 | 1.035 | 1.874 | 1.055 | 0.935 | 0.294 | 3.785 |  |  |  |  |
| not available | 11.725 | 0.000 | 5.242 | 26.225 | 5.676 | 0.312 | 0.196 | 164.437 |  |  |  |  |
| % of time spent on admin work |  |  |  |  |  |  |  |  |  |  |  |  |
| <=50% time | 1.000 |  |  |  | 1.000 |  |  |  |  |  |  |  |
| 50%+ time | 0.686 | 0.001 | 0.554 | 0.849 | 0.710 | 0.512 | 0.254 | 1.980 |  |  |  |  |
| no information | 0.669 | 0.131 | 0.397 | 1.127 | 1.041 | 0.975 | 0.082 | 13.226 |  |  |  |  |
| Drugs are available |  |  |  |  |  |  |  |  |  |  |  |  |
| not available | 1.000 |  |  |  | 1.000 |  |  |  |  |  |  |  |
| available | 1.552 | 0.001 | 1.191 | 2.023 | 1.675 | 0.348 | 0.570 | 4.922 |  |  |  |  |
| Any Stockout |  |  |  |  |  |  |  |  |  |  |  |  |
| Stock-Out | 1.000 |  |  |  | 1.000 |  |  |  |  |  |  |  |
| no Stock-out | 0.581 | 0.001 | 0.417 | 0.809 | 0.540 | 0.439 | 0.113 | 2.571 |  |  |  |  |
| Nurse Age |  |  |  |  |  |  |  |  |  |  |  |  |
| <27 yrs | 1.000 |  |  |  | 1.000 |  |  |  |  |  |  |  |
| 27+yrs | 1.797 | 0.000 | 1.441 | 2.240 | 1.698 | 0.308 | 0.613 | 4.702 |  |  |  |  |
| Nurse experience |  |  |  |  |  |  |  |  |  |  |  |  |
| <35 months | 1.000 |  |  |  | 1.000 |  |  |  |  |  |  |  |
| 35+ months | 0.855 | 0.196 | 0.675 | 1.084 | 0.615 | 0.311 | 0.240 | 1.574 |  |  |  |  |
| OPD per day |  |  |  |  |  |  |  |  |  |  |  |  |
| low | 1.000 |  |  |  | 1.000 |  |  |  |  |  |  |  |
| medium | 0.804 | 0.135 | 0.603 | 1.071 | 1.217 | 0.742 | 0.377 | 3.928 |  |  |  |  |
| high | 0.606 | 0.001 | 0.445 | 0.825 | 0.922 | 0.901 | 0.254 | 3.341 |  |  |  |  |
| _cons | 3.267 | 0.000 | 1.865 | 5.723 | 8.133 | 0.017 | 1.461 | 45.258 | 10.040 | 0.000 | 3.613 | 27.898 |
| Random Part |  |  |  |  |  |  |  |  |  |  |  |  |
| PHC: |  |  |  |  |  |  |  |  |  |  |  |  |
| var( _cons) |  |  |  |  | 1.004 |  | 0.575 | 1.755 | 1.380 |  | 0.794 | 2.399 |
| ICC |  |  |  |  | 0.234 |  | 0.149 | 0.348 | 0.296 |  | 0.194 | 0.422 |
| AIC | 3481.326 |  |  |  | 3083.923 |  |  |  | 3069.479 |  |  |  |
| BIC | 3593.635 |  |  |  | 3202.142 |  |  |  | 3116.767 |  |  |  |

### 2. Appropriateness of prescription (QoC)

We have used two approaches to understand the QoC dimension. <u>First</u>, a latent measure of quality was computed by considering the five important indicators of prescription practice and explored factors attributable to QoC. <u>Second</u>, we evaluated appropriateness of investigation &treatment from the clinical stand point.

#### a. Appropriateness as a measure of facility level variation in Quality of care (QoC)

The QoC index was computed using a latent variable comprising of indicators:(i) % of prescriptions wherein sign &/or symptoms were recorded, (ii) % of prescriptions that had diagnosis (final or provisional) documented, (iii) % of prescriptions wherein diagnostic tests (investigation) were recommended, (iv) % of prescription wherein treatments were recorded (antibiotic and injection), (v) and % of prescriptions wherein follow-up/ referral was documented.

We noted substantial variation in all indicators across PHCs (refer table-11 and figure-4). In order to compute a latent variable, we adjusted the variables by taking deviations from WHO reference value and computed a summative score. The score was further divided into five quintiles to create an ordered variable – QoC index. The index was found consistent with all variables included to construct the index. Subsequently, we compared system and provider level factors across five quintiles of QoC (table-12). It was evident that facilities very good on QoC, had young nurses with doctors either less than 35 yrs or over 65 years of age, spending less time in administrative tasks, and experienced nurses. The facilities that were better on QoC were well-stocked with drugs. Interestingly QoC in prescribing practices were better in PHCs with high OP footfall; due to reduced time availability for providers on administrative activities, better stocked with EDL, and nurses that had spent more time (> 35 months) within the system.

**Table 11:** Variation across facilities on QoC indicators

| Indicators | N | Mean | St.Dev | Min | Pctl(25) | Pctl(75) | Max |
| --- | --- | --- | --- | --- | --- | --- | --- |
| % of pres. sign/symptom mentioned | 31 | 30.4 | 28.5 | 0.0 | 5.2 | 53.6 | 98.1 |
| % of pres. diagnosis mentioned | 31 | 22.6 | 21.5 | 0.0 | 7.0 | 29.5 | 93.0 |
| % of pres. tests recommended | 31 | 5.9 | 7.0 | 0.0 | 0.8 | 6.8 | 33.0 |
| % of pres. <2 medicine prescribed | 31 | 11.9 | 6.9 | 0.0 | 6.8 | 16.3 | 28.2 |
| % of pres. antibiotic prescribed | 31 | 65.3 | 11.5 | 43.1 | 56.4 | 74.7 | 90.1 |
| % of pres. injection prescribed | 31 | 13.9 | 11.2 | 0.0 | 5.0 | 19.4 | 44.6 |
| % of pres. Referral /follow up mentioned | 31 | 2.6 | 6.8 | 0.0 | 0.0 | 1.7 | 35.7 |

**Table 12:** Profile of facilities with different level of quality of care in terms of prescribing practices

|  | v.low | low | moderate | good | v.good | Total |
| --- | --- | --- | --- | --- | --- | --- |
| **Age of doctors** |  |  |  |  |  |  |
| <35 years | 42.9 | 14.3 | 20.0 | 28.6 | 60.0 | 32.3 |
| 35-65 years | 14.3 | 42.9 | 20.0 | 28.6 | 0.0 | 22.6 |
| 65+ years | 28.6 | 28.6 | 60.0 | 28.6 | 40.0 | 35.5 |
| Less time (<50% ) spent on admin task | 42.9 | 28.6 | 80.0 | 71.4 | 80.0 | 58.1 |
| % of prescription signs by the doctor | 51.9 | 58.1 | 64.0 | 50.1 | 75.0 | 58.3 |
| Medicine was available (%) | 42.9 | 14.3 | 60.0 | 57.1 | 60.0 | 45.2 |
| Stock out in past 6 months (%) | 28.6 | 28.6 | 20.0 | 28.6 | 60.0 | 32.3 |
| Age of GNM <27 yrs (%) | 71.4 | 42.9 | 0.0 | 85.7 | 40.0 | 51.6 |
| Work experience of GNM (35+ months) | 28.6 | 85.7 | 80.0 | 85.7 | 80.0 | 71.0 |
| Average OPD per day | 41.0 | 52.8 | 27.4 | 42.1 | 57.1 | 45.7 |
| % Urban | 28.6 | 14.3 | 20.0 | 28.6 | 20.0 | 22.6 |
| **No of PHCs** | **7** | **7** | **5** | **7** | **5** | **31** |

**Figure 4:**
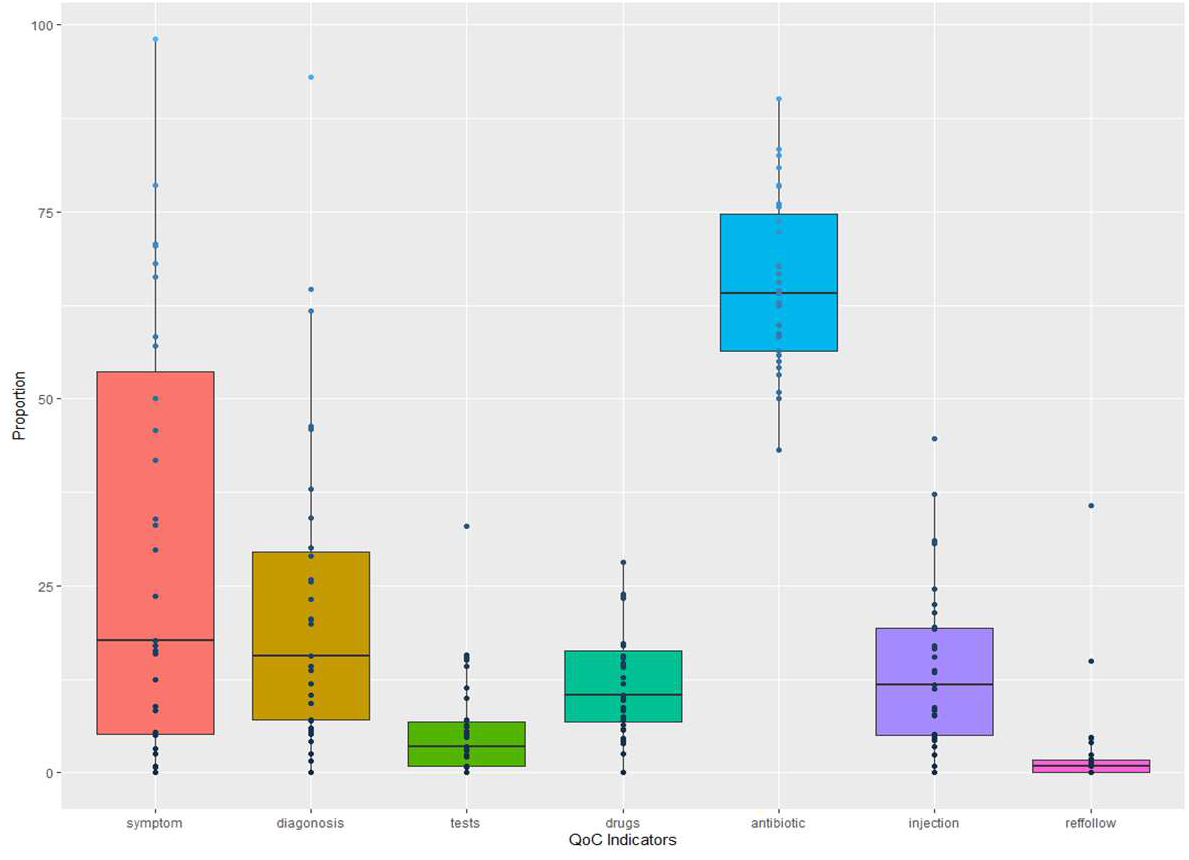
Box plot of QoC indicators

### b. Appropriateness as a measure of Quality of care from clinical stand point

The appropriateness of drug use stems from the Standard Treatment Guidelines (STGs) and best practices to treat certain disease/s. Further, as signs and symptoms are sine-quo-non to patient complaints, from which the clinician draws provisional and differential diagnosis; we analysed 783 prescriptions that had symptoms and signs documented to evaluate appropriateness of drug use. We have used STGs recommended by government of India and WHO to label the appropriateness of prescription. As the study focuses on completeness and compliance of prescription, the diagnosis made by the clinician and the treatment thereof was not questioned. Therefore, the documented justification in terms of investigation advised, treatment given, follow up &referrals advised for such a diagnosis vis-à-vis the documented signs and symptoms was considered relevant for the study, and hence was evaluated.

Nearly 36 % of prescription were found inappropriate in terms of its documented justification for the treatment advised. The commonest inappropriateness in treatment were non-documentation of rationale for:

- Prescription of one or more antibiotics/ 3rd Generation Antibiotic/ Anti-fungal, steroids and NSAIDs, Anti-Rabies vaccine without documentation of signs and symptoms or investigation;
- Using both Gram negative and Gram positive antibiotics for the same event;
- Prescribing Azithromycin in prescription reporting diarrhoea;
- Prescribing steroids for fever and cough in child under five without relevant clinical findings;
- Treatment of dehydration or prescription of fluids in old age without investigating for electrolyte imbalance;
- Not prescribing anti-hypertensive or not advising referral/follow-up for hypertension in pregnancy.

Commonest prescription with limited error on appropriateness were for maternity cases, wherein STG was well defined and adequately communicated to the staff.

While weight is an important criterion for calculation of drug dose, especially in case of children; only in 43 prescriptions weight was documented, which included 6 children under the age group of 5 years. Similarly, Blood pressure was documented only in 4% of cases; commonest of them being: pregnancy and all cases of gastritis, hypertension, weakness, diarrhoea, and body ache in old age patients.

## D. Conclusion

PHCs are the first point of doctor-patient interaction in the public healthcare delivery system. It is not only an entry to the continuum of care cycle in terms of provision of diagnosis, treatment and appropriate referrals and follow-ups; but also echoes with universalization of health care as was envisaged for the first time at Alma-Ata Declaration. Therefore, early diagnosis of disease, its timely treatment and referral happens to be the primary role of a PHC; served through a complete and comprehensive prescription. In order to improve compliance and comprehensiveness of prescription practice in PHC level, our study reveals the need for following system and provider level strengthening efforts:

- **System level Strengthening**
  - Creating facility level enabling environment by ensuring availability of: (i) drugs and supplies, test kits at all times; (ii) uniform prescription format, with sufficient space for recording details as required by MCI; (iii) unique identifier for each patient for maintaining continuum of care across life cycle of the patient; (iv) referral practice to higher centres and back to be streamlined (documented, standardised, and communicated across).
  - Policy reform to rationalise polypharmacy beyond WHO definition, considering Indian realities; thereby including multivitamins as nutritional supplements to all vulnerable sections of the population instead of categorising it as “Drug” in a prescription.
- **Provider level strengthening**
  - Empowering staff nurse /GNM as “Physician Assistant” by protocoled task shifting to treat select diseases with limited allowable prescription &sign off by “Nurse Practitioner” to increase accountability of the system.
  - Stakeholder buy-in to influence behaviour change management, to: (i) improve legibility of prescriptions, and documentation of justification for the treatment offered; (ii) implement Anti- Microbial Stewardship Program to build capacity of care providers on rational drug use.

## Data Availability

All data and information collected is in excel

https://bmcpublichealth.biomedcentral.com/articles/10.1186/s12889-016-3428-8#CR12

## Footnotes

7 For the purpose of the study, legibility of prescription was defined as the effortless ability of reader/ data entry expert to read and copy the contents of the prescription in the content analysis sheet

8 M=Maternity, E=Emergency, P1=Paediatrics of age <5 yrs, P2=Paediatrics of age (>5 yrs and < 15 yrs), Adult (male and female of age between 18-59 yrs of age with conditions other than maternity) and Old Age (=/>60yrs of age)

9 M=Monday, TU=Tuesday, W=Wednesday, TH=Thursday, F=Friday, SA=Saturday

10 The facility/system level factors include: urban /rural location, OPD footfall, availability of medicines, time spent of administrative activities. The provider level factors includes: age and working experience with LEHS|WISH model

11 Ayurveda Yoga Unani Siddha and Homeopathy system of Indian Medicine.

